# High infectiousness immediately before COVID-19 symptom onset highlights the importance of contact tracing

**DOI:** 10.1101/2020.11.20.20235754

**Authors:** W.S. Hart, P.K. Maini, R.N. Thompson

## Abstract

Understanding changes in infectiousness during COVID-19 infections is critical to assess the effectiveness of public health measures such as contact tracing. Data from known source-recipient pairs can be used to estimate the average infectiousness profile of infected individuals, and to evaluate the proportion of presymptomatic transmissions. Here, we infer the infectiousness profile of COVID-19 infections using a mechanistic approach, and show that this method provides an improved fit to data from source-recipient pairs compared to previous studies. Our results indicate a higher proportion of presymptomatic transmissions than previously thought, with many transmissions occurring shortly before symptom onset. High infectiousness immediately prior to symptom onset highlights the importance of contact tracing, even if contacts from a short time window before symptom onset alone are traced.

## Main text

The precise proportion of SARS-CoV-2 transmissions arising from non-symptomatic (either presymptomatic or asymptomatic) infectors remains uncertain (*1, 2*). Statistical models can be used to assess the relative contributions of presymptomatic and symptomatic transmission using data from known source-recipient transmission pairs (*3*–*6*). The distributions of three important epidemiological time periods – the generation time (i.e., the difference between the infection times of the source and recipient) (*3, 4, 7, 8*), the time from onset of symptoms to transmission (TOST) (*4, 9, 10*), and the serial interval (i.e., the difference between the symptom onset times of the source and recipient) (*4, 11*) – can also be inferred. The generation time and TOST distributions also indicate the average infectiousness profile of a host at each time since infection and time since symptom onset, respectively (*9, 12*). This is important for assessing the effectiveness of public health measures such as contact tracing (*3*) and for deciding the duration of quarantine/isolation periods (*13*). Estimates of the generation time distribution of SARS-CoV-2 obtained from transmission pair data have typically involved an assumption that the infectiousness of a host is independent of their symptom status (*3, 7, 8, 14, 15*). However, such an assumption may both be unjustified (*15, 16*) and lead to a poor fit to data (*4*).

Here, we develop a mechanistic approach for inference from transmission pair data. Our method provides an improved fit to data from SARS-CoV-2 transmission pairs compared to previously used approaches, namely: (i) a model assuming that transmission and symptoms are independent (*3, 7, 8, 14*), and (ii) a previous statistical method in which this assumption is relaxed (*4*). We use the fitted models to infer the distributions of epidemiological time intervals and the contribution of presymptomatic infectious individuals to transmission. According to our best-fitting model, we find that the predicted proportion of presymptomatic transmissions is higher than estimated using standard approaches, with a substantial proportion of transmissions occurring in a short time window prior to symptom onset. Finally, we consider the implications of our results for contact tracing.

Transmission pair data (Fig. 1A) generally comprise symptom onset dates for known source-recipient pairs. These data may be supplemented with partial information about infection times, consisting of a range of possible exposure dates for the source and/or recipient (*3*). While the serial interval for each pair can be calculated directly from the data (with some uncertainty, given the unknown precise times of symptom appearance on the onset dates (*17*)), other time intervals, including the generation time and TOST (which is negative for presymptomatic transmissions), are unobserved. In almost all previous approaches that have been used to estimate the generation time distribution of SARS-CoV-2 from transmission pair data (Fig. 1B, left panel), the infectiousness of the source at a given time since infection is assumed to be independent of their incubation period (*3, 7, 8, 14*). In contrast, in our mechanistic approach (Fig. 1B, right panel), which is based on compartmental modelling, each infected host passes through three stages of infection – latent (*E*), presymptomatic infectious (*P*), and symptomatic infectious (*I*). Infectiousness is assumed to be constant during each stage but may vary between presymptomatic and symptomatic infectious hosts.

**Fig. 1.**
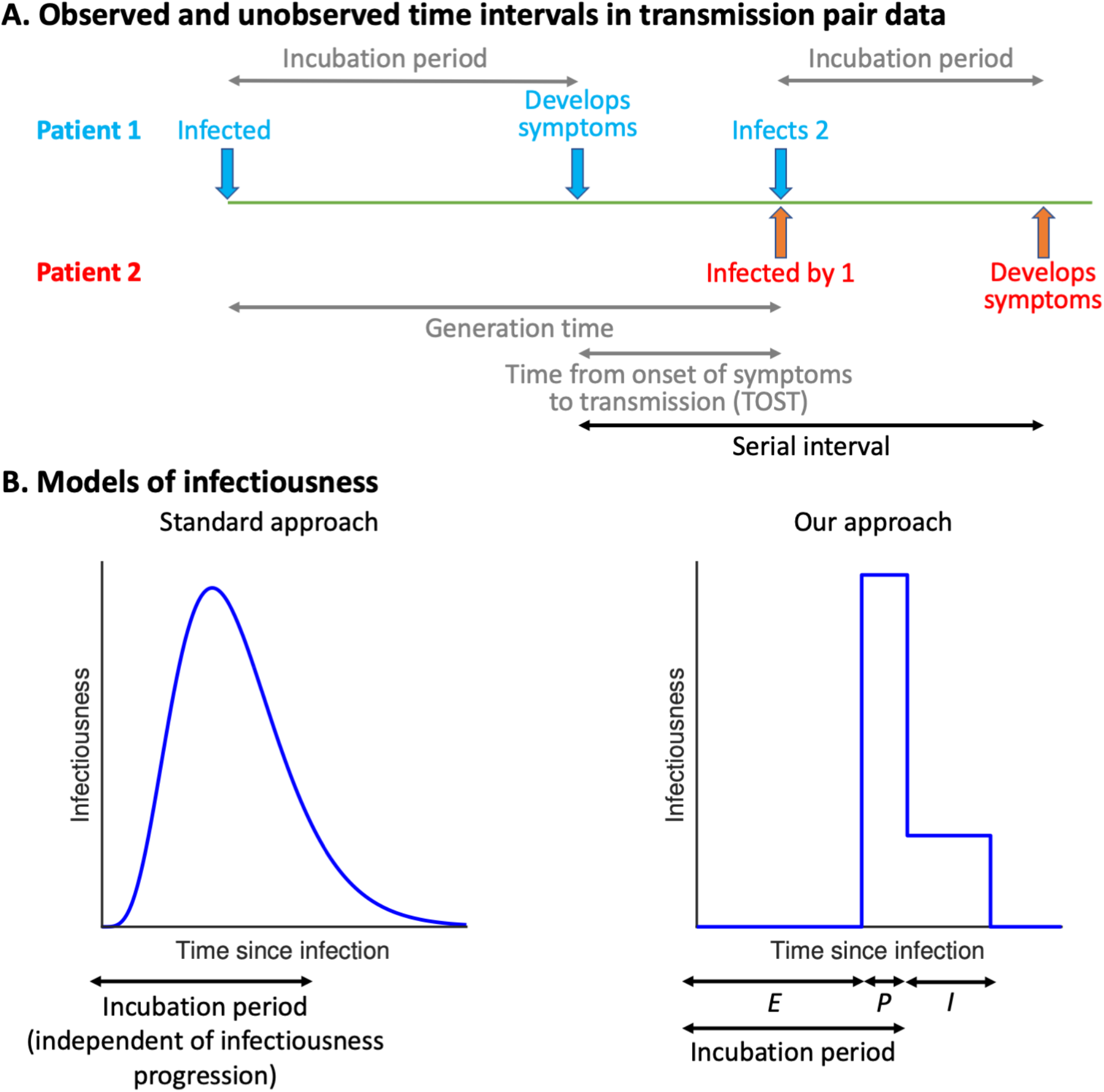
Schematic illustrating time intervals in data from source-recipient pairs and approaches for inference from transmission pair data. A. Important epidemiological time periods in transmission pair data, with time intervals that are not observed directly in grey. B. Assumptions made about the relationship between infectiousness and symptoms within individuals. In standard approaches (left panel), the infectiousness of a host at a given time since infection is independent of their incubation period. In our approach (right panel), we assume that individuals are not infectious during the latent (*E*) period, and that infectiousness may vary between the presymptomatic infectious (*P*) and symptomatic infectious (*I*) periods, for example due to changing behaviour in response to symptoms (*18*).

We considered four different models of infectiousness:

1. Our mechanistic approach, with the relative infectiousness levels for presymptomatic and symptomatic (*I*) infectious hosts estimated from the transmission pair data (the “variable infectiousness model”).
2. Our mechanistic approach, with identical infectiousness levels for presymptomatic\ and symptomatic (*I*) infectious hosts (the “constant infectiousness model”).
3. The best-fitting model from (*4*) (the “Ferretti model”).
4. The standard approach (*3, 8*) in which infectiousness is independent of symptom status (the “independent transmission and symptoms model”).

Following (*4*), we fitted each model to data from 191 SARS-CoV-2 transmission pairs obtained by combining five different datasets (*3, 9, 19*–*21*). To account for uncertainty in the precise times of symptom appearance within the day of onset for the source and recipient (*22*), we used data augmentation Markov chain Monte Carlo (MCMC). The best fit to the data was obtained using our mechanistic approach in which infectiousness varies between presymptomatic infectious and symptomatic hosts (the variable infectiousness model; ΔAIC = 0). The constant infectiousness model gave the next best fit (ΔAIC = 4.5), followed by the Ferretti model (ΔAIC = 8.3). Finally, the standard assumption of independent transmission and symptoms led to a significantly worse fit compared to the other models (ΔAIC = 44.6).

For each model, we calculated the predicted distributions of the generation time (Fig. 2A), TOST (Fig. 2B) and serial interval (Fig. 2C) under the set of model parameters that gave the best fit to the data. The empirical serial interval distribution is also plotted in Fig. 2C, for visual confirmation of the goodness of fit of the different models. We found that the variability in the generation time between individuals was lower for the independent transmission and symptoms model compared to the other three models (Fig. 2A). On the other hand, the TOST distribution was most concentrated around the time of symptom onset nfor the best-fitting variable infectiousness model, and least concentrated for the independent transmission and symptoms model (Fig. 2B). In our best-fitting model, infectiousness was found to decrease immediately after onset, likely due to behavioural factors that reduce the transmission risk following symptom onset (*18*).

**Fig. 2.**
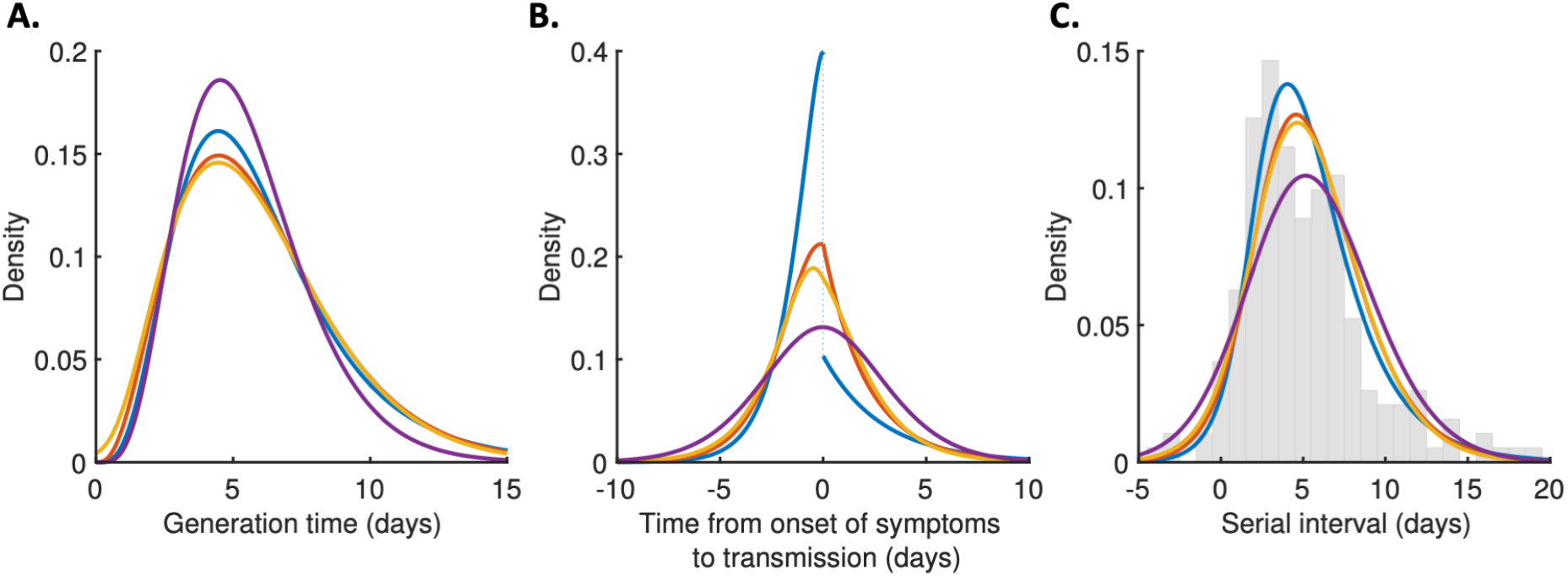
Distributions of epidemiological time intervals estimated by fitting different models to data from 191 SARS-CoV-2 transmission pairs. A. Generation time, indicating the relative expected infectiousness of a host at each time since infection. B. Time from onset of symptoms to transmission (TOST), indicating the relative expected infectiousness of a host at each time since symptom onset. C. Serial interval, indicating the periods between sources and recipients developing symptoms. In panel C, the empirical serial interval distribution from the transmission pair data is shown as grey bars. In addition, discretised versions of the serial interval distributions, calculated using the method in (*23*), are shown in the Supplementary Materials (Fig. S1). In all panels, lines represent: variable infectiousness model (blue), constant infectiousness model (red), Ferretti model (orange), and independent transmission and symptoms model (purple).

Using the posterior distributions of model parameters that were obtained when we fitted the models to data, we calculated the posterior distribution of the proportion of transmissions occurring prior to symptom onset for each model (Fig. 3A). The median (95% CI) presymptomatic proportion was 0.65 (0.53-0.76), 0.56 (0.50-0.62), 0.55 (0.48-0.62), and 0.49 (0.43-0.56) under the variable infectiousness model, constant infectiousness model, Ferretti model, and independent transmission and symptoms model, respectively. Our central estimate of 65% of transmissions occurring prior to symptom onset using our best-fitting model is higher than estimated in previous studies that have fitted statistical models of infectiousness using transmission pair data (*3, 4, 9*) – for example, estimates of 37% and 55% were obtained under an assumption of independent transmission and symptoms in (*3*).

**Fig. 3.**
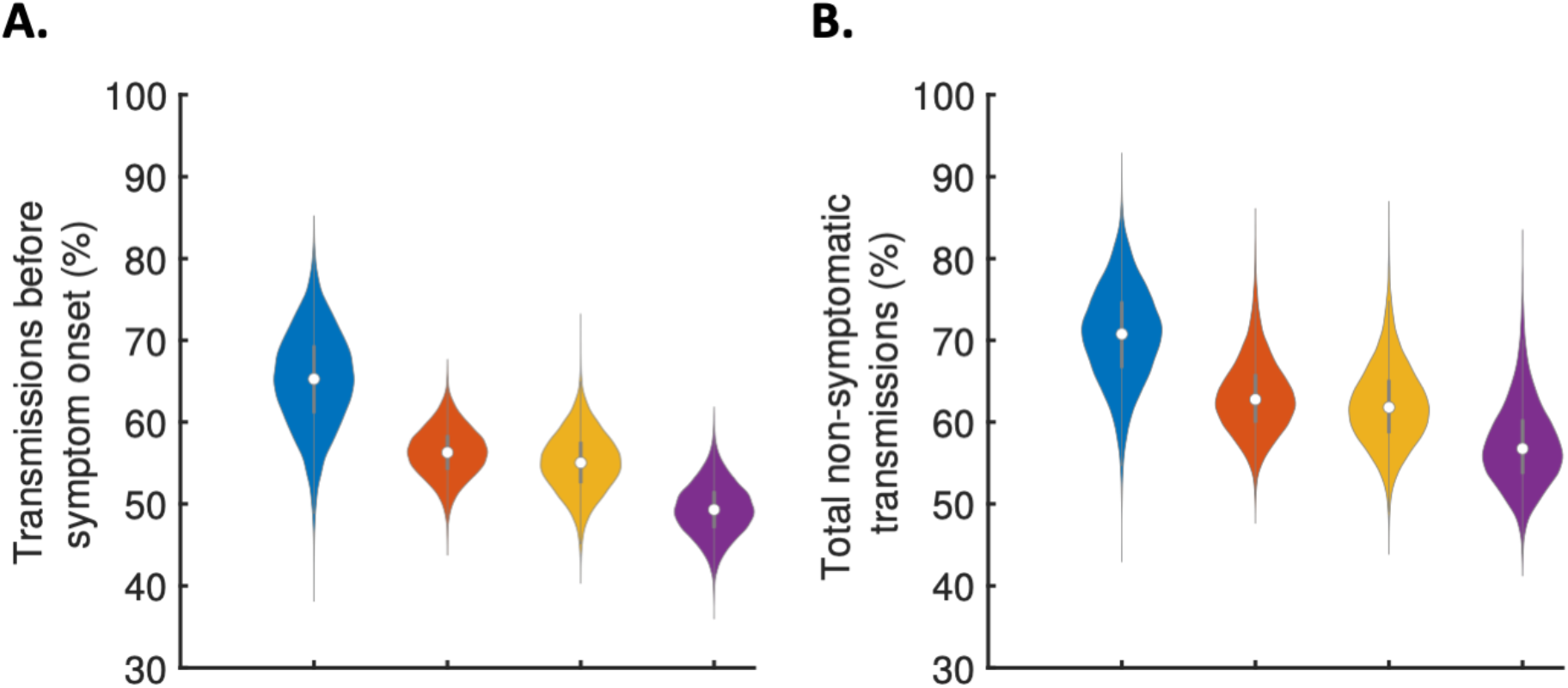
The contribution of non-symptomatic infectious individuals to transmission. A. Violin plots indicating posterior distributions for the proportion of transmissions occurring prior to symptom onset for individuals who develop symptoms (i.e., neglecting transmissions from individuals who remain asymptomatic throughout infection) for the different models. B. Posterior distributions of the total proportion of non-symptomatic transmissions, accounting for transmissions from asymptomatic infectious individuals, for the different models. In both panels, violins represent: variable infectiousness model (blue), constant infectiousness model (red), Ferretti model (orange), and independent transmission and symptoms model (purple).

The estimates in Fig. 3A describe the proportion of transmissions that occur prior to symptom onset, and therefore apply only to individuals who go on to develop symptoms. However, these estimates can be combined with the results of a previous study (*1*) in which the extent of asymptomatic transmission (i.e., transmissions from individuals who never display symptoms) was characterised (Fig. S2), to obtain estimates for the total proportion of non-symptomatic (i.e., either presymptomatic or asymptomatic) transmissions in an entire population of hosts for the different models (Fig. 3B). Again, the non-symptomatic proportion was highest for the variable infectiousness model, and lowest for the independent transmission and symptoms model.

Finally, we considered the implications of these results for contact tracing (Fig. 4). In Fig. 4A, we show the proportion of infectious contacts of hosts who go on to develop symptoms that are identified if contacts are traced up to different times before the symptom onset time of the index case (i.e., for different contact elicitation windows). Long duration contact elicitation windows are impractical and place significant strain on contact tracing systems, leading to contact elicitation windows of two days being used in countries such as the UK and USA (*25*). In the best-fitting variable infectiousness model, 84% of infectious contacts are estimated to be identified when tracing up to 2 days before symptom onset (Fig. 4A, blue dashed). This is due to the clustering of transmission events around the symptom onset time (*cf*. Fig. 2B) and compares to a lower estimate of only 74% if the standard assumption of independence between transmission and symptoms is made (Fig. 4A, purple). We also explored the effect of the timing of isolation of infected individuals identified through contact tracing, and estimated the reduction in onwards transmissions from infected contacts if contact tracing is conducted quickly (Fig. 4B). Compared to the best-fitting variable infectiousness model, the standard independent transmission and symptoms model estimated a similar reduction in transmission when a host was isolated within 4 days of exposure, but under-predicted the efficacy of isolation later in infection. We assumed that contact identification and isolation is perfectly effective in this analysis, but we also explored the sensitivity of our results to this assumption in the Supplementary Materials. In each case that we considered, under the best-fitting model a two-day contact elicitation window is more effective than estimated using existing approaches (Fig. S3).

**Fig. 4.**
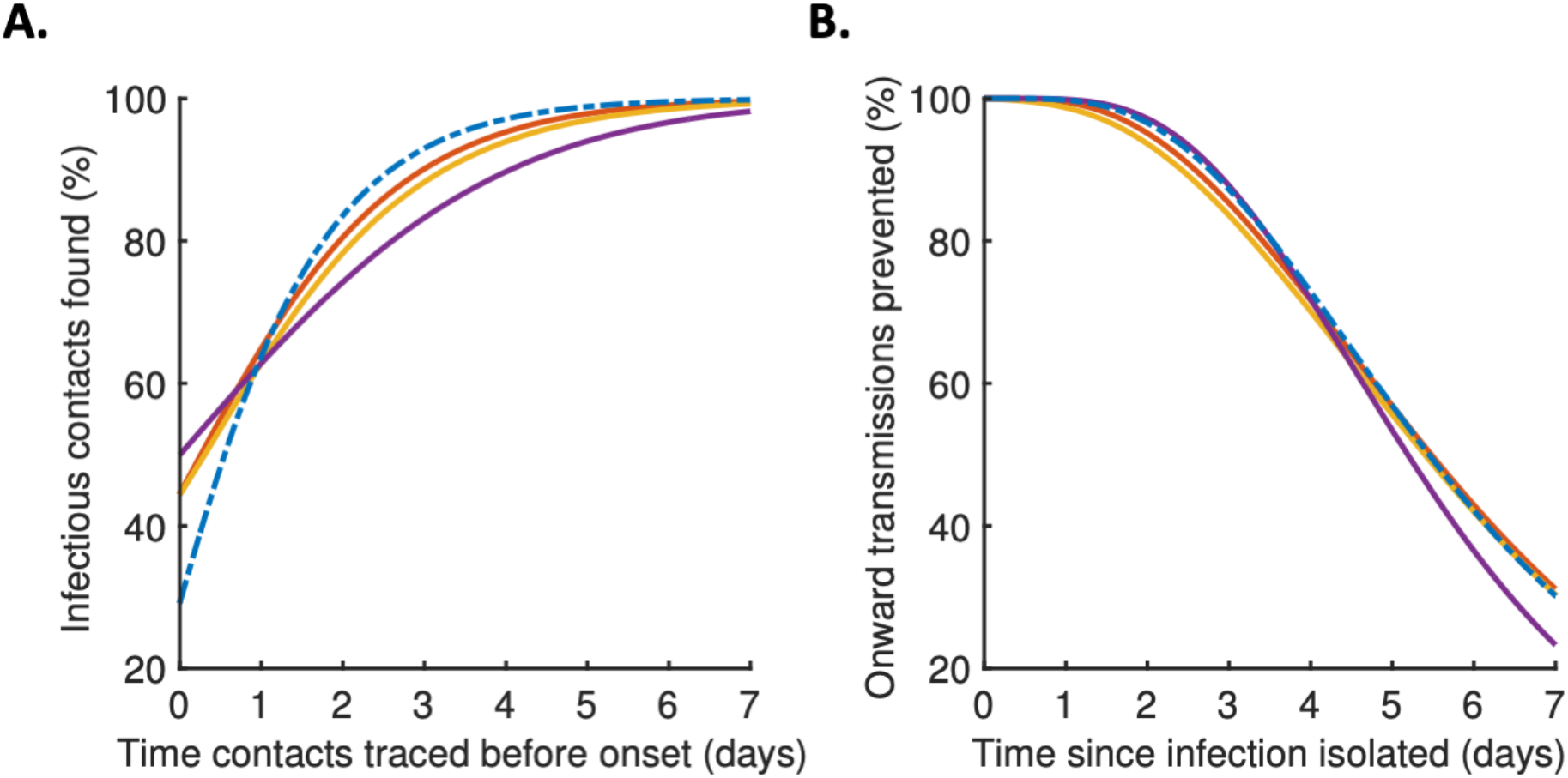
Implications for contact tracing. A. Effect of the contact elicitation window: the proportion of infectious contacts found for different times up to which contacts are traced before the symptom onset time of the index case. B. Effect of the timing of isolation: the proportion of onward transmissions prevented through isolation, for different time periods between exposure and isolation. In both panels, lines represent: variable infectiousness model (blue dashed), constant infectiousness model (red), Ferretti model (orange), and independent transmission and symptoms model (purple).

As we have shown, our mechanistic approach provides an improved fit to transmission pair data, and predicts a higher proportion of transmissions occurring in a short time window before symptom onset, compared to previous approaches. These conclusions are robust to the exact incubation period distribution that we assumed (*26*) when fitting the different models to transmission pair data (Fig. S4). Our best-fitting model outperforms a model predicated on a critical assumption – that infectiousness is independent of symptom status – which underlies most previous studies in which the generation time distribution of COVID-19 is estimated (*3, 7, 8, 14*). That assumption neglects potential relationships between symptoms and viral shedding, as well as behavioural changes in response to symptom onset (*18*). Some alternative assumptions have also been considered when fitting to transmission pair data, such as the possibility that infectiousness depends only on the time since symptom onset, independent of the time of infection (*4, 27*). If the serial interval is always positive, which is not always the case for COVID-19 (*11*), this is equivalent to assuming that the serial interval and generation time distributions are identical (*15, 23, 28*). In a previous study (*4*), another model (the Ferretti model) was developed in which a host’s infectiousness could depend on both the time since infection and the time since symptom onset, and was found to outperform models in which their infectiousness depends on either one of these two times alone. However, as we have demonstrated, our mechanistic approach provides an improved fit to data compared to the statistical Ferretti model. In addition, our method has the advantage of being useful for parameterising population-scale compartmental epidemic forecasting models, since the time periods in our approach correspond naturally to compartments (*29*).

In summary, using a mechanistic approach to infer key epidemiological quantities from transmission pair data indicates that a higher proportion of SARS-CoV-2 transmissions occur prior to symptoms than previously thought. Furthermore, a significant proportion of transmissions arise shortly before symptom onset, indicating that contact tracing is beneficial even if the contact elicitation window is short. Continued use and refinement of contact tracing programmes in countries worldwide is therefore of clear public health importance.

## Materials and Methods

### Notation and general details

Here, we outline the notation used in this section when describing the different models that we considered. For a given transmission pair, we label the source as 1 and the recipient 2, and define:

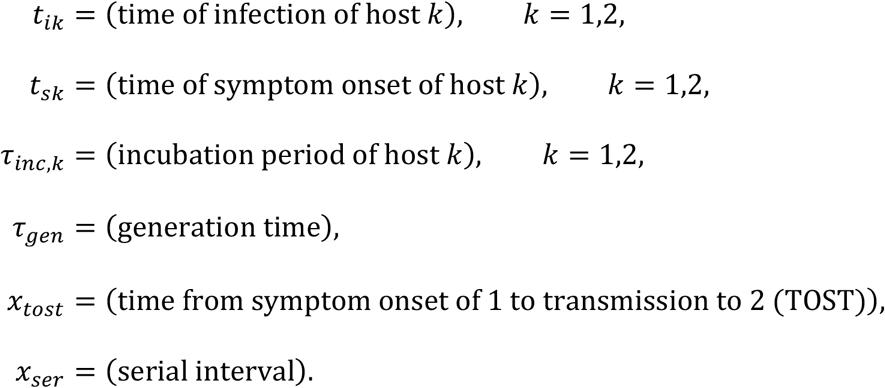

Here, *t* is used to denote calendar times, *τ* for time intervals relative to the time of infection, and *x* for time intervals relative to the time of symptom onset. We denote the probability density functions of the incubation period, generation time, TOST and serial interval as *f*_*inc*_, *f*_*gen*_, *f*_*tost*_ and *f*_*ser*_, respectively, and use a capital *F* for the corresponding cumulative distribution functions.

In addition, we denote the expected infectiousness of a host at time since infection *τ* as *β*(*τ*), and the expected infectiousness at time since symptom onset *x* as *b*(*x*). Note that

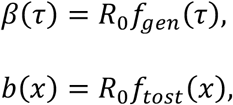

where *R*_*0*_ is the basic reproduction number (for hosts that develop symptoms at some stage during infection). We also let *β*(*τ* | *τ*_*inc*_) and *b*(*x* | *τ*_*inc*_) be the expected infectiousness at time *τ* since infection and at time *x* since onset, respectively, conditional on an incubation period of *τ*_*inc*_ (note that *β*(*τ* | *τ*_*inc*_) = *b*(*τ* − *τ*_*inc*_ | *τ*_*inc*_) and *b*(*x* | *τ*_*inc*_) = *β*(*x* + *τ*_*inc*_ | *τ*_*inc*_)).

We considered several different models for infectiousness (details of individual models are given below). In each model, the conditional infectiousness, *β*(*τ* | *τ*_*inc*_), or equivalently, *b*(*x* | *τ*_*inc*_), is specified. The distributions of the generation time and TOST can be recovered from this conditional infectiousness by averaging over the incubation period distribution (which is assumed to be known):

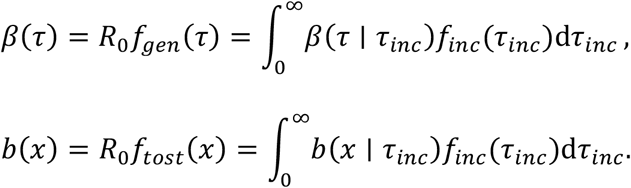

Alternative (equivalent) expressions for the generation time and TOST distributions are available for some of the models considered (these are detailed in the “Models of infectiousness” subsection below).

To obtain an expression for the serial interval distribution, we note that

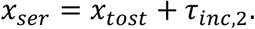

We assume throughout that *x*_*tost*_ and *τ*_*inc*,2_ are independent, so that the serial interval distribution is given by the convolution

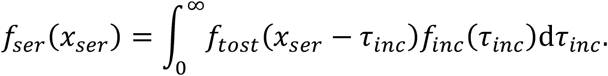

The proportion of presymptomatic transmissions (out of all transmissions generated by individuals who develop symptoms) can be calculated as

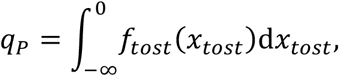

although simpler equivalent expressions for individual models are also detailed later.

#### Data

Following (*4*), we considered COVID-19 transmission pair data from five different studies (*3, 9, 19*–*21*), totalling 191 source-recipient pairs. In all 191 transmission pairs, both the source and the recipient developed symptoms, and the symptom onset date of each host was recorded. In four of the five studies (*3, 9, 19, 20*), intervals of exposure were available for either the source or recipient (or both), whereas in the other (*21*), only symptom onset dates were recorded.

### Incubation period

In the main text, the incubation period was assumed to follow a Gamma distribution with shape parameter 5.807 and scale parameter 0.948 (*26*). This corresponds to a mean incubation period of 5.5 days and a standard deviation of 2.3 days. However, to demonstrate that our main conclusions are robust to the exact incubation period distribution used, we also repeated our analyses using an alternative, more dispersed, Gamma distributed incubation period with a mean of 5.3 days and a standard deviation of 3.2 days (*30*) (Fig. S4).

### Models of infectiousness

### Independent transmission and symptoms model

In this model, the infectiousness of each host at a given time since infection is assumed to be independent of their incubation period, so that

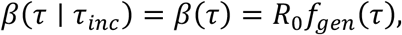

where the generation time distribution, *f*_*gen*_, is prescribed. We assumed (*3, 8*) that

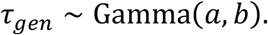

The shape parameter (a) and scale parameter (*b*) were estimated when we fitted the model to transmission pair data.

The TOST distribution for this model is given by

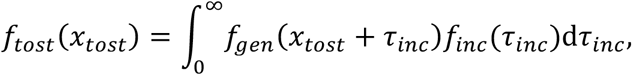

whilst the proportion of presymptomatic transmissions is

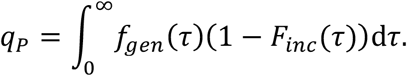

Derivations of these expressions are given in the Supplementary Text.

### Ferretti model

Ferretti et al. (*4*) proposed a model in which the conditional infectiousness was specified as the re-scaled skew-logistic distribution,

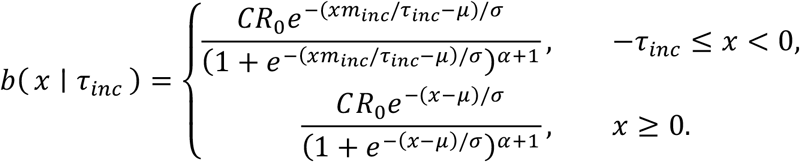

Here, *m*_*inc*_ is the mean incubation period, *μ*, σ and *α* are estimated parameters, and we set

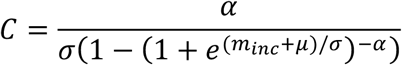

in order to ensure the correct scaling for the infectiousness (see the Supplementary Text).

The proportion of presymptomatic transmissions is

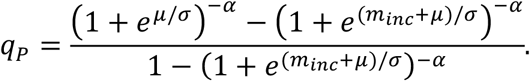

A derivation of this expression is given in the Supplementary Text.

### Our mechanistic model

In our mechanistic approach, we divided each infection into three stages: latent (*E*), presymptomatic infectious (*P*), and symptomatic infectious (*I*). The stage durations were assumed to be independent, and infectiousness was assumed to be constant over the duration of each stage. We denote the stage durations by *y*_*E/P/I*_, their density and cumulative distribution functions by *f*_*E/P/I*_ and *F*_*E/P/I*_, and the infectiousness of hosts in the *P* and *I* stages by *β*_*P/I*_, respectively. We also define the ratio

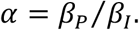

Note that in this model, the basic reproduction number is

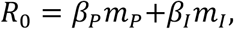

where *m*_*P/I*_ are the respective mean durations of the *P* and *I* stages.

We further assumed that the durations of each stage followed Gamma distributions, with

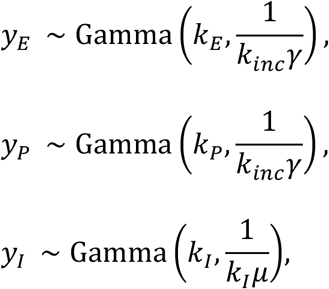

Where

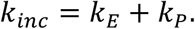

In particular, the scale parameters of *y*_*E*_ and *y*_*P*_ were both assumed to be equal to 1/(*k*_*inc*_*γ*), in order to ensure a Gamma distributed incubation period,

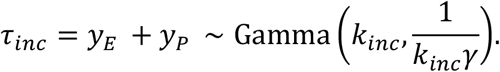

We fixed *k*_*inc*_ = 5.807 and *γ* = 1/(5.807 × 0.948), in order to obtain the specified incubation period distribution (see “Incubation period” subsection above). When we fitted the model to data, we assumed that *k*_*I*_ = 1, so that the symptomatic infectious period followed an exponential distribution. The parameters *k*_*E*_ and *μ* were estimated in the fitting procedure. We considered two versions of the model: one in which we assumed *α* = 1 (the constant infectiousness model), and one in which *α* was also estimated (the variable infectiousness model).

For this model, the infectiousness of a host at time *x* since symptom onset, conditional on an incubation period of *τ*_*inc*_, can be calculated to be

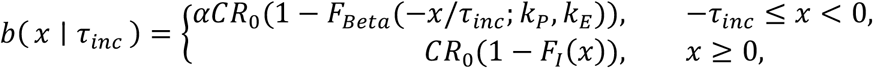

where *F*_*Beta*_(*s*; *a, b*) is the cumulative distribution function of a Beta distributed random variable with parameters *a* and *b*, and

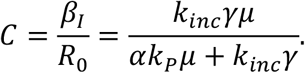

The TOST distribution is given by

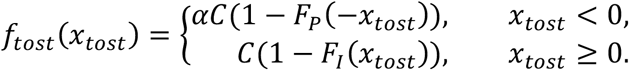

The generation time can be written as

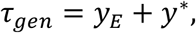

where *y*^*^ is the time between the start of the *P* stage and the transmission occurring, and therefore the generation time distribution is given by the convolution

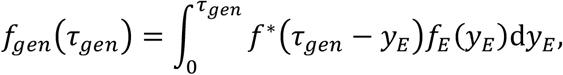

where the density, *f*^*^, of *y*^*^ satisfies

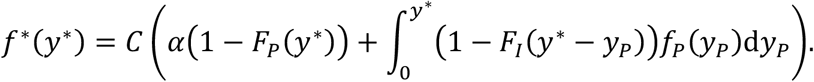

The proportion of presymptomatic transmissions is

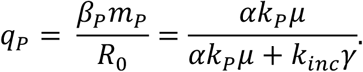

Derivations of these formulae are given in the Supplementary Text.

### Likelihood and model fitting

For a single transmission pair (labelled *n*), suppose that the times of infection for the source and recipient are known to lie in the intervals [*t*_*i*1,*L*_, *t*_*i*1,*R*_] and [*t*_*i2,L*_, *t*_*i*2,*R*_], respectively (where these intervals may be infinite), and that their symptom onset times, *t*_*s*1_ and *t*_*s*2_, are known exactly. In this case, the likelihood of the parameters, *θ*, of the model of infectiousness under consideration (when only that transmission pair is observed) is given by

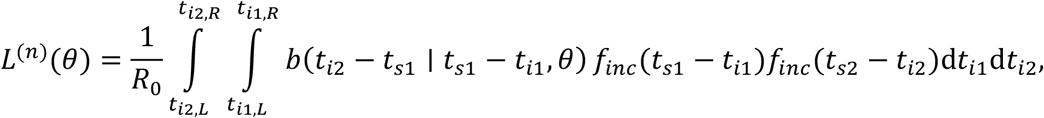

where the dependence of the conditional expected infectiousness, *b*(*x* | *τ*_*inc*_, *θ*), on the model parameters, *θ*, is indicated explicitly. A derivation of this expression is given in the Supplementary Text. Assuming that each transmission pair in our dataset was independent, the overall likelihood was therefore given by the product of the contributions, *L*^*(n*)^(*θ*), from each individual transmission pair, i.e.,

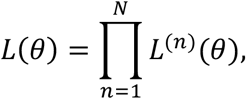

where *N* is the total number of transmission pairs.

In order to account for uncertainty in the exact symptom onset times within the day of onset, we fitted the models to the data using data augmentation MCMC. In particular, in alternating steps of the chain, we updated either the vector of model parameters, *θ*, or the symptom onset times of each source and recipient. The chain was run for 2.5 million steps, of which the first 500,000 were discarded as burn-in. Posterior distributions of parameters were obtained by recording only every 100 iterations of the chain (assuming a uniform prior distribution for each model parameter), whilst the parameter values that maximised the likelihood were taken as point estimates. Full details of the model fitting procedure are given in the Supplementary Text.

### Distributions of the presymptomatic and total non-symptomatic proportion of transmissions

Expressions for the proportion of transmissions, *q*_*P*_, generated prior to symptom onset, are given for the individual models above. Once asymptomatic cases are accounted for, the overall non-symptomatic proportion of transmissions can be written as

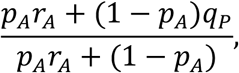

where *p*_*A*_ is the proportion of infected individuals who remain asymptomatic, and *r*_*A*_ is the ratio between the average number of secondary cases generated by an asymptomatic host and the number generated by a host who at some point develops symptoms. A derivation of this expression is given in the Supplementary Text.

For each model, we used the posterior parameter distributions that were obtained when we fitted the model to data to obtain a sample from the posterior distribution of *q*_*P*_. In order to estimate the total proportion of non-symptomatic transmissions, we assumed the distributions

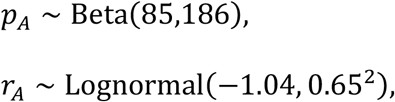

which are consistent with estimates in (*1*). These distributions are shown in Fig. S2. We then combined samples from the assumed distributions of *p*_*A*_ and *r*_*A*_ with the sample that we generated from the posterior distribution of *q*_*P*_ to obtain a distribution for the total proportion of non-symptomatic transmissions.

### Contact tracing

First, we considered the proportion of the infectious contacts of a symptomatic index case that will be found, if contacts are traced up to *d*_1_ days before the time of symptom onset (of the index host). In this case, assuming that it is possible to trace a fraction *ε*_1_ of the host’s total contacts between times −*d*_1_ and ∞ since symptom onset, then the proportion of infectious contacts found is equal to

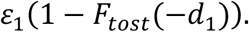

Note that we assumed that the TOST distribution for a detected index case does not differ from that of a random infected individual.

We then considered the proportion of transmissions that can be prevented, if an infected individual (identified through contact tracing) is isolated *d*_2_ days after exposure. Assuming that a proportion *ε*_2_ of infectious contacts that would otherwise occur are prevented during the isolation period, the overall proportion of onward infections prevented through isolation is

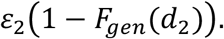

In the main text, we assumed that *ε*_1_ = *ε*_2_ = 1 (i.e., contact identification and isolation are both 100% effective), although values of *ε*_1_ and *ε*_2_ below 1 are considered in the Supplementary Materials (Fig. S3).

## Data Availability

The transmission pair data used in our analyses are available in the Supplementary Materials. Code for reproducing our results is available at https://github.com/will-s-hart/COVID-19-Infectiousness-Profile.

## Acknowledgments

Thanks to members of the Wolfson Centre for Mathematical Biology at the University of Oxford for useful discussions about this work.

## Author contributions

RNT and WSH conceived the research; All authors designed the study; WSH carried out the research; WSH drafted the manuscript; RNT and PKM supervised the research; All authors revised the manuscript and gave final approval for publication.

## References

1. D. Buitrago-Garcia, D. Egli-Gany, M. J. Counotte, S. Hossmann, H. Imeri, A. M. Ipekci, G. Salanti, N. Low, Occurrence and transmission potential of asymptomatic and presymptomatic SARS-CoV-2 infections: A living systematic review and meta-analysis. PLoS Med. 17, e1003346 (2020).

2. M. Casey, J. Griffin, C. G. McAloon, A. W. Byrne, J. M. Madden, D. McEvoy, A. B. Collins, K. Hunt, A. Barber, F. Butler, E. A. Lane, K. O. Brien, P. Wall, K. A. Walsh, S. J. More, Pre-symptomatic transmission of SARS-CoV-2 infection: a secondary analysis using published data. medRxiv (2020), DOI:10.1101/2020.05.08.20094870.

3. L. Ferretti, C. Wymant, M. Kendall, L. Zhao, A. Nurtay, L. Abeler-Dörner, M. Parker, D. Bonsall, C. Fraser, Quantifying SARS-CoV-2 transmission suggests epidemic control with digital contact tracing. Science. 368, eabb6936 (2020).

4. L. Ferretti, A. Ledda, C. Wymant, L. Zhao, V. Ledda, L. Abeler-Dörner, M. Kendall, A. Nurtay, H.-Y. Cheng, T.-C. Ng, H.-H. Lin, R. Hinch, J. Masel, A. M. Kilpatrick, C. Fraser, The timing of COVID-19 transmission. medRxiv (2020), DOI:10.1101/2020.09.04.20188516.

5. W. Zhang, Estimating the presymptomatic transmission of COVID19 using incubation period and serial interval data. medRxiv (2020), DOI:10.1101/2020.04.02.20051318.

6. Y. Liu, Centre for Mathematical Modelling of Infectious Diseases nCoV Working Group, S. Funk, S. Flasche, The contribution of pre-symptomatic infection to the transmission dynamics of COVID-2019. Wellcome Open Res. 5, 58 (2020).

7. Y. Deng, C. You, Y. Liu, J. Qin, X. H. Zhou, Estimation of incubation period and generation time based on observed length-biased epidemic cohort with censoring for COVID-19 outbreak in China. Biometrics, 1–13 (2020).

8. T. Ganyani, C. Kremer, D. Chen, A. Torneri, C. Faes, J. Wallinga, N. Hens, Estimating the generation interval for coronavirus disease (COVID-19) based on symptom onset data, March 2020. Eurosurveillance. 25, 2000257 (2020).

9. X. He, E. H. Y. Lau, P. Wu, X. Deng, J. Wang, X. Hao, Y. C. Lau, J. Y. Wong, Y. Guan, X. Tan, X. Mo, Y. Chen, B. Liao, W. Chen, F. Hu, Q. Zhang, M. Zhong, Y. Wu, L. Zhao, F. Zhang, B. J. Cowling, F. Li, G. M. Leung, Temporal dynamics in viral shedding and transmissibility of COVID-19. Nat. Med. 26, 672–675 (2020).

10. P. Ashcroft, J. S. Huisman, S. Lehtinen, J. A. Bouman, C. L. Althaus, R. R. Regoes, S. Bonhoeffer, COVID-19 infectivity profile correction. Swiss Med. Wkly. 150, w20336 (2020).

11. Z. Du, X. Xu, Y. Wu, L. Wang, B. J. Cowling, L. A. Meyers, Serial interval of COVID-19 among publicly reported confirmed cases. Emerg. Infect. Dis. 26, 1341–1343 (2020).

12. C. Fraser, Estimating individual and household reproduction numbers in an emerging epidemic. PLoS One. 2, e758 (2007).

13. P. Ashcroft, S. Lehtinen, S. Bonhoeffer, Quantifying the impact of quarantine duration on COVID-19 transmission. medRxiv (2020), DOI:10.1101/2020.09.24.20201061.

14. J. Knight, S. Mishra, Estimating effective reproduction number using generation time versus serial interval, with application to covid-19 in the Greater Toronto Area, Canada. Infect. Dis. Model. 5, 889–896 (2020).

15. S. Lehtinen, P. Ashcroft, S. Bonhoeffer, On the relationship between serial interval, infectiousness profile and generation time. medRxiv (2020), DOI:10.1101/2020.09.18.20197210.

16. S. Bacallado, Q. Zhao, N. Ju, Letter to the editor: Generation interval for COVID-19 based on symptom onset data. Eurosurveillance. 25, 2001381 (2020).

17. R. N. Thompson, J. E. Stockwin, R. D. van Gaalen, J. A. Polonsky, Z. N. Kamvar, P. A. Demarsh, E. Dahlqwist, S. Li, E. Miguel, T. Jombart, J. Lessler, S. Cauchemez, A. Cori, Improved inference of time-varying reproduction numbers during infectious disease outbreaks. Epidemics. 29, 100356 (2019).

18. P. Manfredi, A. D’Onofrio, Eds., Modeling the interplay between human behavior and the spread of infectious diseases (Springer, New York, 2013).

19. W. Xia, J. Liao, C. Li, Y. Li, X. Qian, X. Sun, H. Xu, G. Mahai, X. Zhao, L. Shi, J. Liu, L. Yu, M. Wang, Q. Wang, A. Namat, Y. Li, J. Qu, Q. Liu, X. Lin, S. Cao, S. Huan, J. Xiao, F. Ruan, H. Wang, Q. Xu, X. Ding, X. Fang, F. Qiu, J. Ma, Y. Zhang, A. Wang, Y. Xing, S. Xu, Transmission of corona virus disease 2019 during the incubation period may lead to a quarantine loophole. medRxiv (2020), DOI:10.1101/2020.03.06.20031955.

20. H. Y. Cheng, S. W. Jian, D. P. Liu, T. C. Ng, W. T. Huang, H. H. Lin, Contact Tracing Assessment of COVID-19 Transmission Dynamics in Taiwan and Risk at Different Exposure Periods before and after Symptom Onset. JAMA Intern. Med. 180, 1156– 17 1163 (2020).

21. J. Zhang, M. Litvinova, W. Wang, Y. Wang, X. Deng, X. Chen, M. Li, W. Zheng, L. Yi, X. Chen, Q. Wu, Y. Liang, X. Wang, J. Yang, K. Sun, I. M. Longini, M. E. Halloran, P. Wu, B. J. Cowling, S. Merler, C. Viboud, A. Vespignani, M. Ajelli, H. Yu, Evolving epidemiology and transmission dynamics of coronavirus disease 2019 outside Hubei province, China: a descriptive and modelling study. Lancet Infect. Dis. 23 20, 793–802 (2020).

22. R. N. Thompson, Novel coronavirus outbreak in Wuhan, China, 2020: intense surveillance is vital for preventing sustained transmission in new locations. J. Clin. Med. 9, 498 (2020).

23. A. Cori, N. M. Ferguson, C. Fraser, S. Cauchemez, A new framework and software to estimate time-varying reproduction numbers during epidemics. Am. J. Epidemiol. 178, 1505–1512 (2013).

24. NHS test and trace: how it works, (Available at https://www.gov.uk/guidance/nhs-test-and-trace-how-it-works).

25. Investigating a COVID-19 Case, (Available at https://www.cdc.gov/coronavirus/2019-ncov/php/contact-tracing/contact-tracing-plan/investigating-covid-19-case.html).

26. S. A. Lauer, K. H. Grantz, Q. Bi, F. K. Jones, Q. Zheng, H. R. Meredith, A. S. Azman, N. G. Reich, J. Lessler, The incubation period of coronavirus disease 2019 (CoVID-19) from publicly reported confirmed cases: estimation and application. Ann. Intern. Med. 172, 577–582 (2020).

27. E. L. Davis, T. C. D. Lucas, A. Borlase, T. M. Pollington, S. Abbott, D. Ayabina, T. Crellen, J. Hellewell, L. Pi, G. F. Medley, T. D. Hollingsworth, P. Klepac, An imperfect tool: COVID-19 “test & trace” success relies on minimising the impact of false negatives and continuation of physical distancing. medRxiv (2020), DOI:10.1101/2020.06.09.20124008.

28. T. Britton, G. S. Tomba, Estimation in emerging epidemics: Biases and remedies. J. R. Soc. Interface. 16, 20180670 (2019).

29. W. S. Hart, P. K. Maini, C. A. Yates, R. N. Thompson, A theoretical framework for transitioning from patient-level to population-scale epidemiological dynamics: influenza A as a case study. J. R. Soc. Interface. 17, 20200230 (2020).

30. N. Linton, T. Kobayashi, Y. Yang, K. Hayashi, A. Akhmetzhanov, S. Jung, B. Yuan, R. Kinoshita, H. Nishiura, Incubation period and other epidemiological characteristics of 2019 novel coronavirus infections with right truncation: a statistical analysis of publicly Available case data. J. Clin. Med. 9, 538 (2020).

