## Supplementary Materials for "High infectiousness immediately before COVID-19 symptom onset highlights the importance of contact tracing"

#### Supplementary Text

Derivation of the likelihood

For a given transmission pair, the joint probability density that:

i. patient 1 (the source) is infected in the time interval $[t_{i1,L}, t_{i1,R}]$;

ii. patient 1 transmits the pathogen to patient 2 (we write $1 \to 2$ to denote the transmission occurring) in the time interval $[t_{i2,L}, t_{i2,R}]$;

and

iii. patients 1 and 2 develop symptoms at times $t_{s1}$ and $t_{s2}$, respectively;

conditioned on the parameters, $\theta$, of the model of infectiousness under consideration, is given by

$$p\left(1 \to 2, t_{s1}, t_{s2}, [t_{i1,L}, t_{i1,R}], [t_{i2,L}, t_{i2,R}] \mid \theta\right)$$

$$= \int_{t_{i2,L}}^{t_{i2,R}} \int_{t_{i1,L}}^{t_{i1,R}} p(1 \to 2, t_{i1}, t_{s1}, t_{i2}, t_{s2} \mid \theta)\, \mathrm{d}t_{i1} \mathrm{d}t_{i2}$$

$$= \int_{t_{i2,L}}^{t_{i2,R}} \int_{t_{i1,L}}^{t_{i1,R}} p(1 \to 2, t_{i2}, t_{s2} \mid t_{i1}, t_{s1}, \theta) p(t_{i1}, t_{s1} \mid \theta)\, \mathrm{d}t_{i1} \mathrm{d}t_{i2}$$

$$= \int_{t_{i2,L}}^{t_{i2,R}} \int_{t_{i1,L}}^{t_{i1,R}} p(t_{s2} \mid 1 \to 2, t_{i1}, t_{s1}, t_{i2}, \theta) p(1 \to 2, t_{i2} \mid t_{i1}, t_{s1}, \theta) p(t_{i1}, t_{s1} \mid \theta)\, \mathrm{d}t_{i1} \mathrm{d}t_{i2}$$

$$= \int_{t_{i2,L}}^{t_{i2,R}} \int_{t_{i1,L}}^{t_{i1,R}} p(1 \to 2, t_{i2} \mid t_{i1}, t_{s1}, \theta) p(t_{s1} \mid t_{i1}, \theta) p(t_{i1} \mid \theta)\, p(t_{s2} \mid t_{i2}, \theta) \mathrm{d}t_{i1} \mathrm{d}t_{i2}.$$

We note that

$$p(1 \to 2, t_{i2} \mid t_{i1}, t_{s1}, \theta) \propto b(t_{i2} - t_{s1} \mid t_{s1} - t_{i1}, \theta).$$

This is because the left-hand side gives the probability of a transmission from 1 to 2 occurring at time $t_{i2}$, conditioned on the infection and onset times of 1, and is therefore proportional to the conditional infectiousness, $b(x_{tost} \mid \tau_{inc}, \theta)$. We also have that

$$p(t_{sk} \mid t_{ik}, \theta) = f_{inc}(t_{sk} - t_{ik}),$$

for $k = 1,2$. In an exponentially growing epidemic with growth rate $r$, the term $p(t_{i1} \mid \theta)$ will introduce a factor proportional to $e^{rt_{i1}}$ into the likelihood (*1*), although we neglect this correction here (note that we found a similar fit to data using the Ferretti model compared to that obtained in (*2*), in which the same model was fitted to the same dataset but including this correction). We therefore obtain the expression for the likelihood, $L^{(n)}(\theta)$, given in Materials and Methods, up to a constant scaling factor. The factor $1/R_0$ was added for convenience, although note that in general,

$$\frac{1}{R_0}\int_0^{\infty} b(x_{tost} \mid \tau_{inc}, \theta)\mathrm{d}x_{tost}$$

may not be equal to 1, since the average number of secondary infections generated by a host may depend on their incubation period.

Details of model fitting procedure

We denote the vector of model parameters for the model of infectiousness under consideration by $\theta$, the vector of symptom onset times (for each source and recipient) by $\boldsymbol{t}$, and the corresponding likelihood by $L(\theta; \boldsymbol{t})$. We define proposal distributions $Q_1(\theta_{prop} \mid \theta)$ and $Q_2(\boldsymbol{t}_{prop} \mid \boldsymbol{t})$, which are taken to be symmetric (i.e., $Q_1(\theta_{prop} \mid \theta) = Q_1(\theta \mid \theta_{prop})$ and $Q_2(\boldsymbol{t}_{prop} \mid \boldsymbol{t}) = Q_2(\boldsymbol{t} \mid \boldsymbol{t}_{prop})$; the exact proposal distributions we used are detailed below).

The data augmentation MCMC algorithm that we used is given by the following steps:

1. Initialise $\theta = \theta_0$ and $\boldsymbol{t} = \boldsymbol{t}_0$.
2. Calculate $L_0 = L(\theta_0; \boldsymbol{t}_0)$
3. For $m = 1, \dots, M$:
   - If $m$ is odd, sample $\theta_{prop}$ from $Q_1(\theta_{prop} \mid \theta_{m-1})$, and set $\boldsymbol{t}_{prop} = \boldsymbol{t}_{m-1}$.
   - If $m$ is even, sample $\boldsymbol{t}_{prop}$ from $Q_2(\boldsymbol{t}_{prop} \mid \boldsymbol{t}_{m-1})$, and set $\theta_{prop} = \theta_{m-1}$.
   - Calculate $L_{prop} = L(\theta_{prop}; \boldsymbol{t}_{prop})$.
   - Generate a random number, $r$, uniformly distributed between 0 and 1.
   - If $r \leq L_{prop}/L_{m-1}$, set $\theta_m = \theta_{prop}$, $\boldsymbol{t}_m = \boldsymbol{t}_{prop}$ and $L_m = L_{prop}$. Otherwise, set $\theta_m = \theta_{m-1}$, $\boldsymbol{t}_m = \boldsymbol{t}_{m-1}$ and $L_m = L_{m-1}$.

We constrained the symptom onset time, $t_s$, of each host to lie on the grid

$$[t_{s,L} + \delta t, d + 2\delta t, \dots, t_{s,L} + 1],$$

where $t_{s,L}$ is the start of the day of onset for that host, and we took $\delta t = 0.125$ days. The contribution to the likelihood from each transmission pair, $L^{(n)}(\theta)$, was calculated by discretising the integrals (see the "Likelihood and model fitting" subsection in Materials and Methods), with the infection time of a given host, $t_i$, constrained to the grid

$$\left[t_{i,L} + \frac{\delta t}{2}, \dots, t_{i,R} - \frac{\delta t}{2}\right],$$

where $t_{i,L}$ and $t_{i,R}$ are upper/lower bounds for the infection time of that host. Note that different discretisations were used for the infection and onset times, both to avoid conditioning on an incubation period of zero days (since the conditional infectiousness may be undefined in this case) and to avoid the possibility of transmissions occurring at the exact time of symptom onset (since the infectiousness profile was allowed to be discontinuous at the onset time in our mechanistic model). We also assumed a maximum possible incubation period of 30 days.

For each model we considered, the initial parameter values, $\theta_0$, were chosen arbitrarily. The initial symptom onset times, $\boldsymbol{t}_0$, were uniformly and independently sampled on the grid of possible onset times for each host. Independent proposal distributions were used for each model parameter. For parameters, $\theta^{(j)}$, which could either be positive or negative, we used a normal proposal distribution, whereas for positive parameters we used a lognormal proposal distribution. In particular, we set

$$\theta^{(j)}_{prop} = \theta^{(j)}_{current} + r,$$

for parameters that could take positive or negative values, and

$$\theta^{(j)}_{prop} = \theta^{(j)}_{current} \times e^{r},$$

for positive parameters, where in both cases, $r$ is a normally distributed random variate with mean zero and standard deviation $\sigma^{(j)}$. We also used independent proposal distributions for each symptom onset time, $t_s$, choosing

$$t_{s,prop} = \begin{cases} t_{s,current} - \delta t, & \text{with probability } (1-p)/2, \\ t_{s,current}, & \text{with probability } p, \\ t_{s,current} + \delta t, & \text{with probability } (1-p)/2, \end{cases}$$

where any proposed onset times lying outside the grid of possible values were reset to their previous value. The tuning parameters of these proposal distributions, $\sigma^{(i)}$ and $p$, were chosen to ensure an acceptance rate of between 20% and 30%.

### Model-specific derivations

#### *Independent transmission and symptoms model*

For the independent transmission and symptoms model, the TOST distribution is given by

$$f_{tost}(x_{tost}) = \frac{1}{R_0}\int_0^{\infty} b(x_{tost} \mid \tau_{inc}) f_{inc}(\tau_{inc}) \mathrm{d}\tau_{inc}$$

$$= \frac{1}{R_0}\int_0^\infty \beta(x_{tost}+\tau_{inc} \mid \tau_{inc}) f_{inc}(\tau_{inc})\mathrm{d}\tau_{inc}$$

$$= \int_0^\infty f_{gen}(x_{tost}+\tau_{inc}) f_{inc}(\tau_{inc})\mathrm{d}\tau_{inc}\,.$$

Alternatively, this formula can be derived by noting that

$$x_{tost} = \tau_{gen} - \tau_{inc,1}.$$

In this model, $\tau_{gen}$ and $\tau_{inc,1}$ are assumed to be independent, so the TOST distribution is therefore given by the convolution of the distributions of $\tau_{gen}$ and $-\tau_{inc,1}$.

The proportion of presymptomatic transmissions is given by

$$q_P = \int_{-\infty}^{0} f_{tost}(x_{tost})\mathrm{d}x_{tost}$$

$$= \int_{-\infty}^{0}\int_0^\infty f_{gen}(x_{tost}+\tau_{inc}) f_{inc}(\tau_{inc})\mathrm{d}\tau_{inc}\,\mathrm{d}x_{tost}$$

$$= \int_0^\infty \int_{\tau_{gen}}^\infty f_{gen}(\tau_{gen}) f_{inc}(\tau_{inc}) d\tau_{inc}\,\mathrm{d}\tau_{gen}$$

$$= \int_0^\infty f_{gen}(\tau_{gen})\big(1 - F_{inc}(\tau_{gen})\big)\mathrm{d}\tau_{gen}.$$

*Ferretti model*

To derive the correct scaling factor, $C$, in the conditional infectiousness, we note that we require

$$\int_{-\infty}^{\infty} f_{tost}(x)\mathrm{d}x = \int_{-\infty}^{\infty}\int_0^\infty \frac{1}{R_0} b(x \mid \tau_{inc}) f_{inc}(\tau_{inc}) d\tau_{inc}\,\mathrm{d}x = 1.$$

Now, we can calculate

$$\int_{-\infty}^{\infty} \frac{1}{R_0} b(x \mid \tau_{inc})\mathrm{d}x$$

$$= \int_{-\tau_{inc}}^{0} \frac{Ce^{-(xm_{inc}/\tau_{inc}-\mu)/\sigma}}{(1+e^{-(xm_{inc}/\tau_{inc}-\mu)/\sigma})^{\alpha+1}} \mathrm{d}x + \int_{0}^{\infty} \frac{Ce^{-(x-\mu)/\sigma}}{(1+e^{-(x-\mu)/\sigma})^{\alpha+1}} \mathrm{d}x$$

$$= \frac{C\sigma}{\alpha}\left[1-\left(1+e^{\mu/\sigma}\right)^{-\alpha} + \frac{\tau_{inc}}{m_{inc}}\left(\left(1+e^{\mu/\sigma}\right)^{-\alpha} - \left(1+e^{(m_{inc}+\mu)/\sigma}\right)^{-\alpha}\right)\right].$$

Therefore,

$$\int_{-\infty}^{\infty}\int_{0}^{\infty} \frac{1}{R_0} b(x \mid \tau_{inc}) f_{inc}(\tau_{inc}) \mathrm{d}\tau_{inc}\, \mathrm{d}x$$

$$= \int_{0}^{\infty}\left(\int_{-\infty}^{\infty} \frac{1}{R_0} b(x \mid \tau_{inc}) \mathrm{d}x\right) f_{inc}(\tau_{inc}) \mathrm{d}\tau_{inc}$$

$$= \frac{C\sigma}{\alpha}\left[1-\left(1+e^{(m_{inc}+\mu)/\sigma}\right)^{-\alpha}\right] = 1,$$

so we have

$$C = \frac{\alpha}{\sigma(1-(1+e^{(m_{inc}+\mu)/\sigma})^{-\alpha})}.$$

The proportion of presymptomatic transmissions is given by

$$q_P = \int_{-\infty}^{0} f_{tost}(x) \mathrm{d}x$$

$$= \int_{0}^{\infty}\int_{-\infty}^{0} \frac{1}{R_0} b(x \mid \tau_{inc}) f_{inc}(\tau_{inc}) \mathrm{d}x\, \mathrm{d}\tau_{inc}$$

$$= \int_{0}^{\infty} \frac{C\sigma\tau_{inc}}{\alpha m_{inc}}\left[\left(1+e^{\mu/\sigma}\right)^{-\alpha} - \left(1+e^{(m_{inc}+\mu)/\sigma}\right)^{-\alpha}\right] f_{inc}(\tau_{inc}) \mathrm{d}\tau_{inc}$$

$$= \frac{\left(1+e^{\mu/\sigma}\right)^{-\alpha} - \left(1+e^{(m_{inc}+\mu)/\sigma}\right)^{-\alpha}}{1-(1+e^{(m_{inc}+\mu)/\sigma})^{-\alpha}}.$$

*Our mechanistic model*

In our mechanistic model, the expected infectiousness of a host at time $x$ since symptom onset is given by

$$b(x) = \begin{cases} \beta_P \times p(Y_P \geq -x), & x < 0, \\ \beta_I \times p(Y_I \geq x), & x \geq 0, \end{cases}$$

where we here explicitly distinguish the random variables $Y_{E/P/I}$ from their observed values $y_{E/P/I}$ (i.e., the lengths of each stage of infection). Therefore,

$$f_{tost}(x_{tost}) = \frac{1}{R_0} b(x_{tost}) = \begin{cases} \alpha C(1 - F_P(-x_{tost})), & x_{tost} < 0, \\ C(1 - F_I(x_{tost})), & x_{tost} \geq 0, \end{cases}$$

where

$$C = \frac{\beta_I}{R_0} = \frac{\beta_I}{\beta_P m_P + \beta_I m_I} = \frac{1}{\alpha m_P + m_I} = \frac{1}{\left(\frac{\alpha k_P}{k_{inc}\gamma} + \frac{1}{\mu}\right)} = \frac{k_{inc}\gamma\mu}{\alpha k_P \mu + k_{inc}\gamma}.$$

Conditional on an incubation period of length $\tau_{inc}$, the expected infectiousness is

$$b(x \mid \tau_{inc}) = \begin{cases} \beta_P \times p(Y_P \geq -x \mid Y_E + Y_P = \tau_{inc}), & -\tau_{inc} \leq x < 0, \\ \beta_I \times p(Y_I \geq x), & x \geq 0. \end{cases}$$

Now,

$$p(Y_P \geq -x \mid Y_E + Y_P = \tau_{inc}) = \int_{-x}^{\infty} f(Y_P = y_P \mid Y_E + Y_P = \tau_{inc}) \mathrm{d}y_P$$

$$= \int_{-x}^{\infty} \frac{f(Y_E + Y_P = \tau_{inc} \mid Y_P = y_P) f(Y_P = y_P)}{f(Y_E + Y_P = \tau_{inc})} \mathrm{d}y_P$$

$$= \int_{-x}^{\infty} \frac{f_E(\tau_{inc} - y_P) f_P(y_P)}{f_{inc}(\tau_{inc})} \mathrm{d}y_P,$$

where we used Bayes' rule to obtain the second equality. For the special case of Gamma distributed stage durations considered, we have that

$$\frac{f_E(\tau_{inc} - y_P) f_P(y_P)}{f_{inc}(\tau_{inc})} = \frac{1}{\tau_{inc}} f_{Beta}(y_P/\tau_{inc}; k_P, k_E)\,),$$

where $f_{Beta}(x; a, b)$ is the probability density function of a Beta distributed random variable with parameters $a$ and $b$. Therefore,

$$p(Y_P \geq -x \mid Y_E + Y_P = \tau_{inc}) = F_{Beta}(-x/\tau_{inc}; k_P, k_E)\,),$$

and so

$$b(x \mid \tau_{inc}) = \begin{cases} \alpha C R_0 (1 - F_{Beta}(-x/\tau_{inc}; k_P, k_E)), & -\tau_{inc} \le x < 0, \\ C R_0 (1 - F_I(x)), & x \ge 0. \end{cases}$$

The expected infectiousness at time $y^*$ since the start of the *P* stage is equal to

$$b^*(y^*) = \beta_P \times p(Y_P \ge y^*) + \beta_I \times p(Y_P \le y^*, Y_P + Y_I \ge y^*).$$

The second probability can be evaluated by conditioning on the value of $Y_P$, in order to obtain

$$b^*(y^*) = \beta_P\big(1 - F_P(y^*)\big) + \beta_I \int_0^\infty p\big(Y_P \le y^*, Y_P + Y_I \ge y^* \mid Y_P = y_p\big) f_P(y_P) \mathrm{d}y_P$$

$$= \beta_P\big(1 - F_P(y^*)\big) + \beta_I \int_0^{y^*} p\big(Y_I \ge y^* - y_P \mid Y_P = y_p\big) f_P(y_P) \mathrm{d}y_P$$

$$= \beta_P\big(1 - F_P(y^*)\big) + \beta_I \int_0^{y^*} \big(1 - F_I(y^* - y_P)\big) f_P(y_P) \mathrm{d}y_P.$$

Therefore, the distribution of the time between the start of the *P* stage and secondary transmission occurring is

$$f^*(y^*) = C\left(\alpha\big(1 - F_P(y^*)\big) + \int_0^{y^*} \big(1 - F_I(y^* - y_P)\big) f_P(y_P) \mathrm{d}y_P\right).$$

The proportion of presymptomatic transmissions is

$$q_P = \frac{\beta_P m_P}{R_0} = \frac{\beta_P m_P}{\beta_P m_P + \beta_I m_I} = \frac{\alpha m_P}{\alpha m_P + m_I} = \frac{\left(\frac{\alpha k_P}{k_{inc}\gamma}\right)}{\left(\frac{\alpha k_P}{k_{inc}\gamma} + \frac{1}{\mu}\right)} = \frac{\alpha k_P \mu}{\alpha k_P \mu + k_{inc}\gamma}.$$

<u>Derivation of expression for the total proportion of non-symptomatic transmissions once asymptomatic cases are accounted for</u>

The basic reproduction number can be decomposed as

$$R_0 = p_A R_A + (1 - p_A)(R_P + R_I),$$

where $p_A$ is the proportion of completely asymptomatic cases, $R_A$ is the expected number of secondary transmissions generated by each asymptomatic host, and $R_{P/I}$ are the expected numbers of transmissions generated before and after symptom onset by a host who develops symptoms, respectively. The total proportion of non-symptomatic transmissions is given by

$$\frac{p_A R_A + (1 - p_A) R_P}{R_0} = \frac{p_A R_A + (1 - p_A) R_P}{p_A R_A + (1 - p_A)(R_P + R_I)}$$

$$= \frac{p_A x_A + (1 - p_A) q_P}{p_A x_A + (1 - p_A)},$$

where

$$q_P = \frac{R_P}{R_P + R_I}$$

is the proportion of transmissions generated prior to symptom onset by hosts who develop symptoms, and

$$x_A = \frac{R_A}{R_P + R_I}$$

is the ratio between the expected number of transmissions generated by an asymptomatic host, and that generated by a host who develops symptoms.

### Supplementary Figures

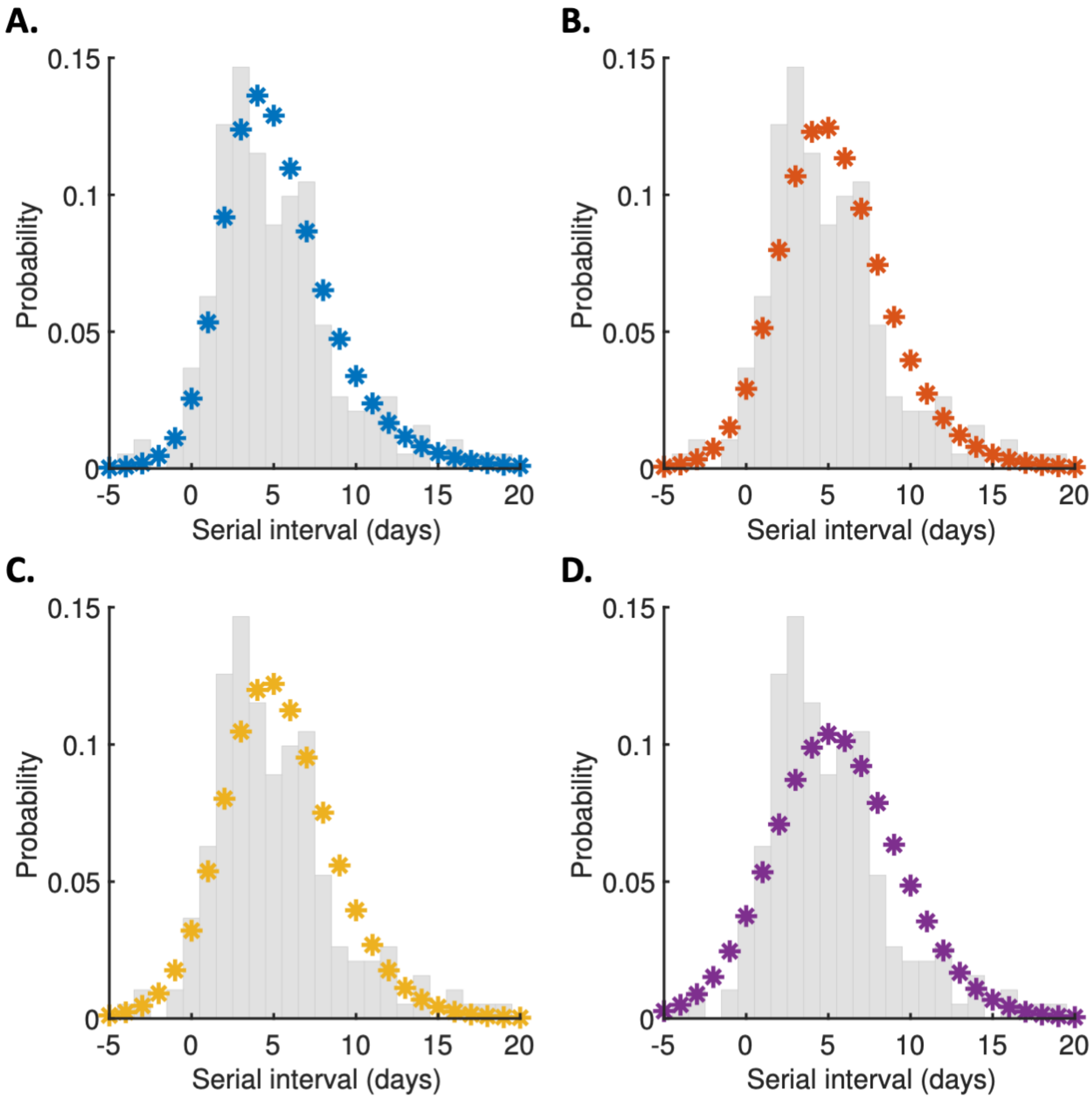


**Fig. S1. Discretised serial interval distributions.** Discretised versions of the serial interval distributions shown in Fig. 2C, calculated using the method in (*3*), plotted as stars alongside the empirical serial interval distribution from the transmission pair data (grey bars). A. Variable infectiousness model. B. Constant infectiousness model. C. Ferretti model. D. Independent transmission and symptoms model.

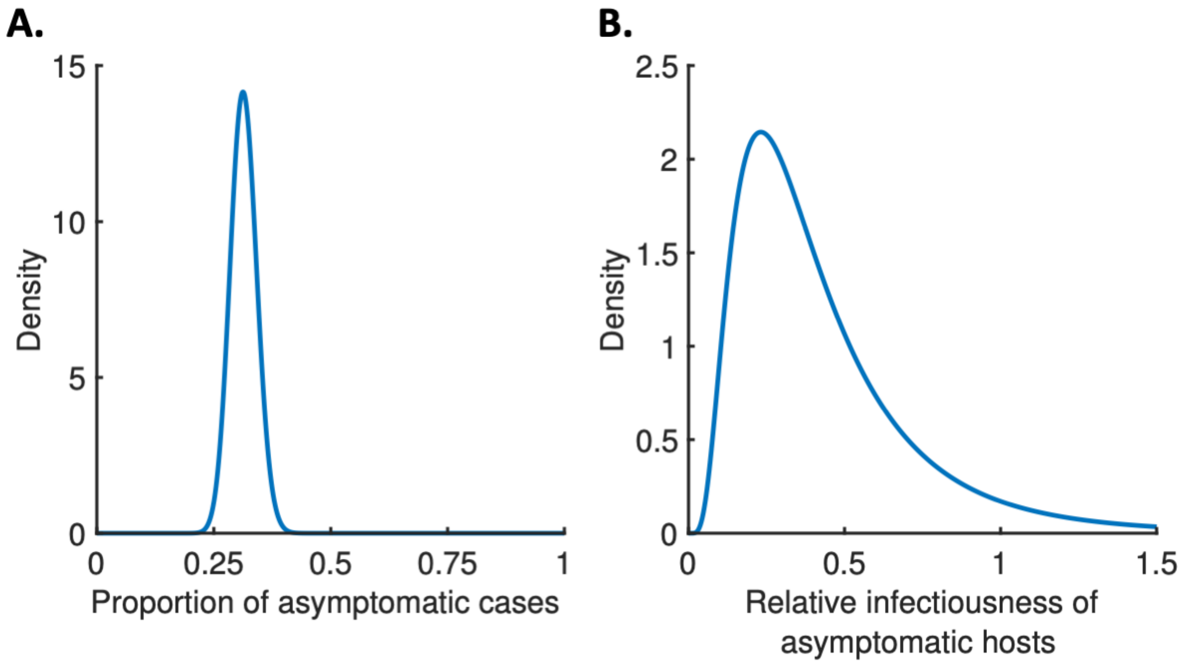


**Fig. S2. The contribution of asymptomatic cases to transmission.** A. Assumed distribution for the proportion of asymptomatic cases, $p_A$. B. Assumed distribution for the relative infectiousness of asymptomatic hosts, $x_A$. Details of how these distributions were obtained are given in Materials and Methods.

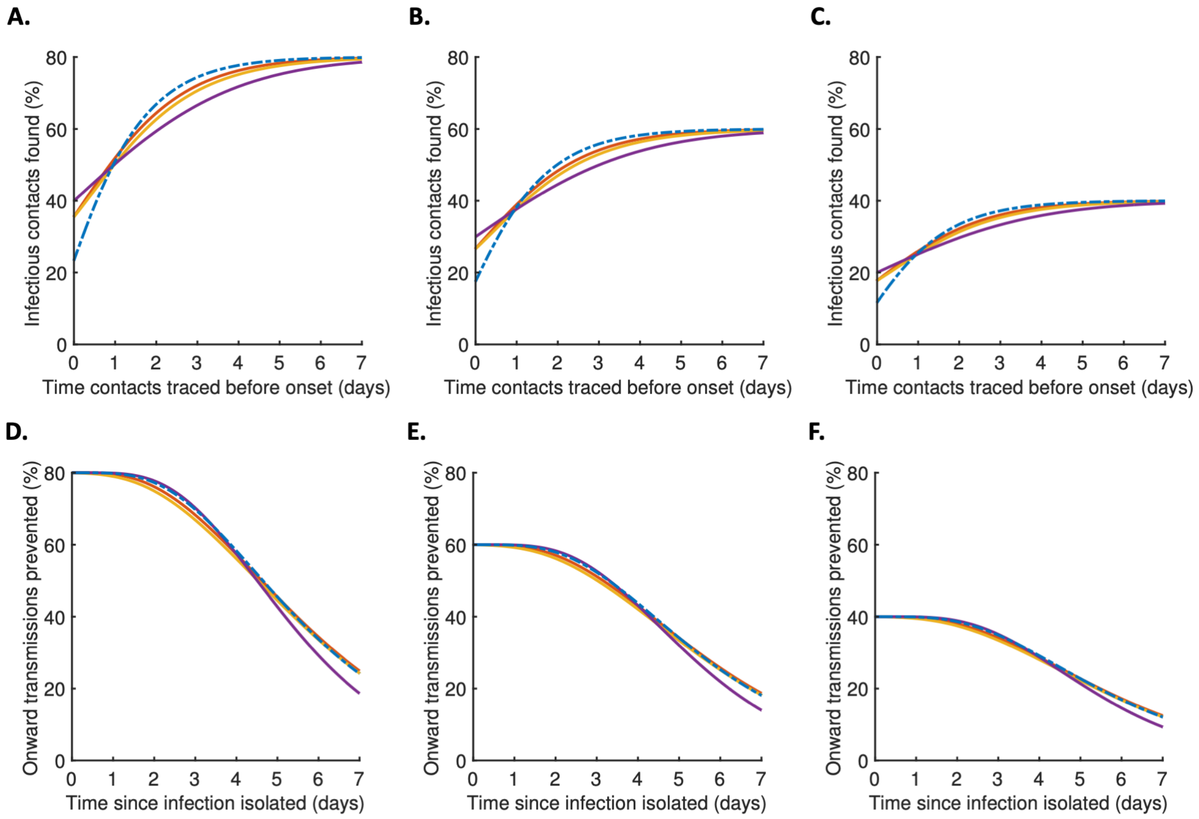


**Fig. S3. Robustness to efficiency of contact tracing and isolation.** A–C. Equivalent panels to Fig. 4A in the main text, for different values of the contact identification effectiveness, $\varepsilon_1$ (see Materials and Methods). A. $\varepsilon_1 = 0.8$. B. $\varepsilon_1 = 0.6$. C. $\varepsilon_1 = 0.4$. D–F. Equivalent panels to Fig. 4B in the main text, for different values of the isolation effectiveness, $\varepsilon_2$ (see Materials and Methods). D. $\varepsilon_2 = 0.8$. E. $\varepsilon_2 = 0.6$. F. $\varepsilon_2 = 0.4$. In all panels, lines represent: variable infectiousness model (blue dashed), constant infectiousness model (red), Ferretti model (orange), and independent transmission and symptoms model (purple).

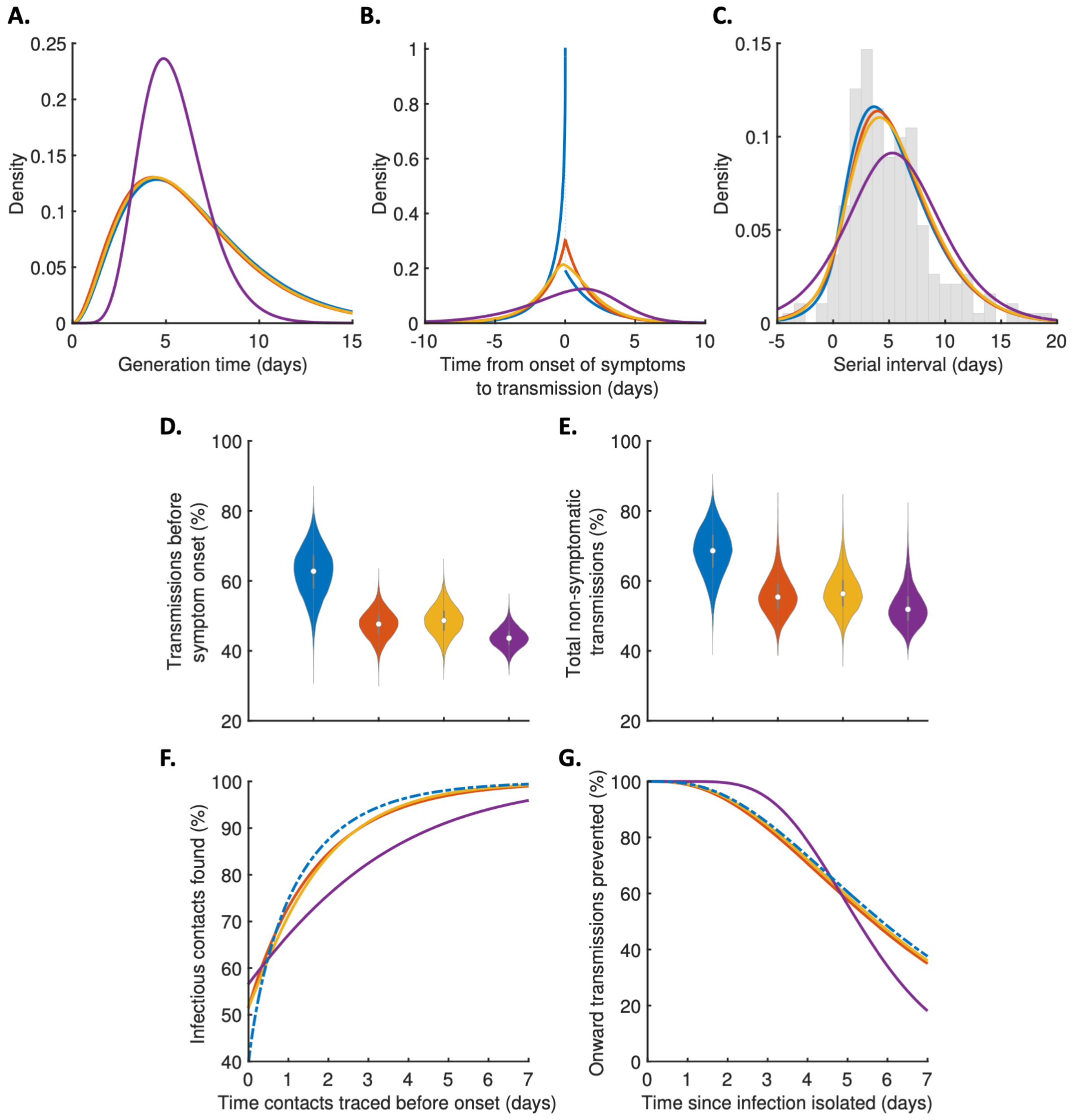


**Fig. S4. Robustness to incubation period distribution.** Equivalent panels to Fig. 2 (A–C), Fig. 3 (D–E) and Fig. 4 (F–G) in the main text, for an alternative incubation period distribution (*4*) (see Materials and Methods). $\Delta$AIC values for the different models are 0 (variable infectiousness model), 7.3 (constant infectiousness model), 13.5 (Ferretti model), 69.2 (independent transmission and symptoms model). In all panels, lines represent: variable infectiousness model (blue, dashed in panels F–G), constant infectiousness model (red), Ferretti model (orange), and independent transmission and symptoms model (purple).

**Table S1.**

Values of fitted parameters for each model. Definitions of the model parameters are given in Materials and Methods.

| Model | Parameter | Point estimate | 95% CI |
|---|---|---|---|
| Variable infectiousness | $k_E$ | 3.93 | 2.45-4.20 |
| | $\mu$ | 0.35 | 0.26-0.53 |
| | $\alpha$ | 3.86 | 0.87-5.96 |
| Constant infectiousness | $k_E$ | 3.05 | 2.08-3.48 |
| | $\mu$ | 0.48 | 0.36-0.58 |
| Ferretti | $\mu$ | −2.65 | (−10.68)-(−0.60) |
| | $\sigma$ | 1.67 | 1.43-2.24 |
| | $\alpha$ | 3.15 | 1.21-112.29 |
| Independent transmission and symptoms | $a$ | 5.63 | 3.73-9.17 |
| | $b$ | 0.98 | 0.60-1.49 |

### References:


1. L. Ferretti, C. Wymant, M. Kendall, L. Zhao, A. Nurtay, L. Abeler-Dörner, M. Parker, D. Bonsall, C. Fraser, Quantifying SARS-CoV-2 transmission suggests epidemic control with digital contact tracing. *Science*. **368**, eabb6936 (2020).
2. L. Ferretti, A. Ledda, C. Wymant, L. Zhao, V. Ledda, L. Abeler-Dörner, M. Kendall, A. Nurtay, H.-Y. Cheng, T.-C. Ng, H.-H. Lin, R. Hinch, J. Masel, A. M. Kilpatrick, C. Fraser, The timing of COVID-19 transmission. *medRxiv* (2020), doi:10.1101/2020.09.04.20188516.
3. A. Cori, N. M. Ferguson, C. Fraser, S. Cauchemez, A new framework and software to estimate time-varying reproduction numbers during epidemics. *Am. J. Epidemiol.* **178**, 1505–1512 (2013).
4. N. Linton, T. Kobayashi, Y. Yang, K. Hayashi, A. Akhmetzhanov, S. Jung, B. Yuan, R. Kinoshita, H. Nishiura, Incubation period and other epidemiological characteristics of 2019 novel coronavirus infections with right truncation: a statistical analysis of publicly available case data. *J. Clin. Med.* **9**, 538 (2020).
